# Measuring Childhood Trauma among Adults in the Health and Retirement Study

**DOI:** 10.64898/2026.01.27.26344534

**Authors:** Adrienne R.S. Lee, David R. Strong, Gretchen E. Bandoli, Linda K. McEvoy, Scott C. Roesch, Eyal Oren, Andrea Z. LaCroix

## Abstract

**Background:** Early life social determinants of health, such as childhood trauma, have implication on adverse health outcomes later in the life course. Our objective was to develop a childhood trauma measure within the Health and Retirement Study (HRS) – a large, diverse, U.S.-based aging cohort.

**Methods:** Data from the HRS Psychosocial and Lifestyle Questionnaire [2006-2016] and Life History Survey [2015-2017]) surveys collected thirteen binary items measuring self-reported exposure to early life adversity across the two study questionnaires. Participants who completed both questionnaires and had exposure items available were included in the analyses. Frequencies and percentages for self-reported trauma items are presented for the study sample and by gender and race/ethnicity. Using complete cases, exploratory factor analyses followed by *Mokken* scale analyses were performed to evaluate the scalability of the childhood trauma items. Predictive criterion validity of the final domains was evaluated with general health and socioeconomic indicators at participant baseline.

**Results:** Among the sample with complete childhood trauma data available (n=9,340), most were women (60.7%), White/Non-Hispanic (73.2%), and had a high school/general education degree (54.0%). The most reported childhood traumas were paternal separation ≥6-months (22.8%), parental death (21.4%), sibling death (18.1%), and problematic parental substance use (17.5%). Two scales were formed based on factor analysis and scalability coefficients. The domain measuring disruption of family structure had strong scalability (*H*_*T*_ = 0.55) and included living in an orphanage, foster care, parents divorced/separated, ≥6-month from mother and/or father, and grandparents as primary caretakers. A second domain measuring adverse experiences of parent and/or sibling death had moderate scalability (*H*_*T*_ = 0.41). Parental substance abuse and physical abuse clustered together in a third domain with weak scalability (*H*_*T*_ = 0.39).

**Conclusions:** The early adversity items available in the HRS offer meaningful domains for which researchers can evaluate childhood trauma exposure in the context of aging outcomes in older adults. In particular, the family structure domain and parental/sibling death demonstrated moderate-to-strong scalability and may have important implications for health trajectories later in life.

## Introduction

Adverse childhood experiences (ACEs), or the exposure to potentially traumatic events before age 18, have lasting implications throughout the life course.^1^ Over 60% of people living in the U.S. have experienced at least one type of ACE, and experiencing multiple ACEs is more prevalent among women and racial/ethnic minorities.^2^ ACEs have a strong, graded, dose response with several adverse health (i.e., smoking, coronary heart disease, diabetes)^1,3^ and social (i.e., less educational attainment, unemployment, household poverty)^4^ outcomes in adulthood.

The National Institute on Aging (NIA): Strategic Directions for Research, 2020-2025, Goal B-3 prioritized future work to identify early life exposures that may differentially impact individual aging trajectories. Childhood socioeconomic adversity is a contemporaneous and distinct exposure from ACEs,^5,6^ that often gets grouped together with psychological trauma/adversity in measures and analyses. However, researchers^5,6^ and the NIA encourage distinguishing childhood trauma from childhood socioeconomic adversity as they are distinct and can modify each other. As such, understanding associations between childhood trauma – independently from childhood socioeconomic adversity – with various aging outcomes, the causal pathways that are operating, and the role of resilience in these relationships may be a key facet to addressing root causes of aging health disparities.

Despite the importance of studying associations between childhood trauma and aging outcomes that has been underscored in recent years, availability of trauma measures within aging cohorts is still limited. The Health and Retirement Study (HRS) is one of the largest and longest-standing cohort studies of older Americans and given extensive available data and linkages, the cohort is well-suited for evaluating the life course impact of ACEs on aging outcomes. To our knowledge, two groups have operationalized ACEs in the HRS, both of which included early life socioeconomic adversity in their measures, and neither of which subjected their measures to psychometric analysis.^7,8^ The objective of this study was to construct a reliable ACE measure(s) of key health-related domains and assess construct validity using available HRS data.

## Methods

### Study Population

The analysis was conducted using data from the University of Michigan’s HRS funded by the NIA and the U.S. Social Security Administration. The HRS is a large, diverse, longitudinal panel study representative of U.S. adults aged ≥50 with data collection waves occurring biennially. Enrollment for the study started in 1992, and a steady-state design replenishes the cohort every six years to maintain representativeness of the population aged ≥50. Stratified multi-stage area probability design with geographic stratification and clustering was used to recruit households, and Black and Hispanic/Latino households were oversampled at twice the rate of White households. At least one age-eligible adult (aged ≥50) was enrolled into the study along with their spouse or other household member(s) regardless of their age. Eligible participants must have completed both the Life History Survey (LHS; 2015, 2017) and the Psychosocial and Lifestyle Questionnaire (PLQ; 2006-2016).

### Measures

#### Childhood trauma

Self-reported experiences of childhood adversity were collected in two mailed and self-administered questionnaires with binary response options. The LHS was administered from 2015-2017 to three subgroups under various conditions (methods reported elsewhere) and ultimately covered all HRS participants in the study as of 2016. The purpose of the questionnaire was to collect information about residential, education, employment and health history, and other childhood and family events. Nine binary items from LHS measured familial adversity before the age of 16 and were used in the subsequent analyses. Participants were asked whether they (1) ever lived in a children’s home or orphanage, (2) ever lived with a foster family/in a foster home, (3) ever lived at a boarding school, (4) if their biological/adoptive parents were separated/divorced, (5) ever were separated from their mother for ≥6 months, (6) ever were separated from their father for ≥6 months, (7) the death of a parent, (8) the death of a sibling, and last, (9) if their grandparent(s) were ever their primary caregiver.

The PLQ was given to a rotating random 50% of participants who received enhanced face-to-face interview in 2004-2016 study waves. The purpose of the questionnaire was to measure participants’ evaluations of their life circumstances, subjective wellbeing, and lifestyle. There were four questions asked from the PLQ about potentially harmful parental behaviors and child behaviors before the age of 18 which were used in the subsequent analyses. Participants were asked whether they (1) ever did a year of school over again, (2) if they were ever in trouble with the police, (3) if their parent(s) ever drank or used drugs so often that it caused problems in the family, and (4) if they were ever physically abused by either of their parents.

#### Subgroups of Interest

Participants self-reported race (White/Caucasian, Black/African American, American Indian, Asian or Other) and Hispanic ethnicity (Mexican, Other, unknown, or non-Hispanic), from which we derived a race/ethnicity variable: White/Non-Hispanic, Black, Hispanic, and Other. Participants also reported their gender (male or female).

#### Validity Measures

To examine predictive construct validity, we assessed correlations between the domains of the final ACE measure(s) with various health indicators in adulthood at the participant’s respective baseline visit. Participants self-reported their general health (excellent, very good, good, fair, or poor); if they ever smoked cigarettes and whether they currently smoked now; if they drink alcohol, on average how many drinks they have in a day (less than one/day, 1-2 drinks/day, 3-4 drinks/day, or 5+ drinks/day), which was dichotomized at >2 drinks/day; whether during the past 12 months there was ever a time they felt sad, blue, or depressed for ≥ two weeks in a row; and last, self-reported weight (in pounds) and height (in feet and inches) were converted to kilograms and meters, and used to estimate body mass index (BMI). Adult educational attainment was also examined as a socioeconomic indicator in adulthood (No degree, HS/GED, and two-year college or higher). Last, we measured childhood financial adversity^7^ (before age 16) using four financial indicators: relative financial status (pretty well off, about average, or poor), father unemployed for several months or more, family moved due to financial difficulty, and the number of books at home.^7^ We summed the items (range: 0–4) and categorized them into no (score: 0), low (score: 1-2), and high (score: 3-4) childhood financial adversity.

### Statistical Analyses

We initially conducted factor analysis and parallel analysis to extract the optimal number of factors underlying the ACE item responses using polychoric correlation matrices as the estimation procedure. In preliminary analyses, a qualitative assessment informed by communalities (ℎ^2^ ≥ 0.3), loading factors (*λ* ≥ 0.4), Cronbach’s alpha (*α*) and omega hierarchical (*ω_h_*) was conducted; to facilitate interpretation, factors with three items and strong loading factors were considered. Following these analyses, *Mokken* Scale Analysis (MSA) was utilized to investigate potential scalability of smaller (< 3 item) subdomains using the ACE items.^9,10^

MSA is a nonparametric Item Response Theory (IRT) model that examines observable item properties and model fitness to obtain sets of items with reasonable discriminatory power.^9^ Assumptions from this MSA model include (1) unidimensionality, (2) local independence, (3) latent monotonicity, and (4) non-intersection.^11^ The *Mokken* R package was used for analyses. First to partition items into *Mokken* scales, automated item selection procedure (AISP), an exploratory hierarchical clustering algorithmic method, was used to examine properties implied by the double monotonicity model. Scalable items were partitioned into *Mokken* scales using a positive lower bound inter-item covariance, *c,* set to 0.3 where item scalability coefficients (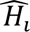) were positive and ≥*c*. Domains were created from partitioned items in the AISP, and item (*H*_*i*_), item-pair (*H*_*i*j_) and test (*H*_*T*_) scalability coefficients were evaluated for fitness. Test scalability coefficients were considered weak if 0.3 ≤ *H*_*T*_ < 0.4, moderate if 0.4 ≤ *H*_*T*_ < 0.5, and strong if *H*_*T*_ ≥ 0.5 relative to the degree to which ordering of individuals by test score reflects the ordering of items on the latent trait, *θ*. Latent monotonicity (R function: **check.monotonicity**) and non-intersection (R functions: **check.pmatrix** and **check.restscore**) assumptions were investigated. A default minimum violation (*minvi)* of *m*=0.03 was used to avoid statistical testing of small violations of non-intersection. An alpha level of 0.05 was used to assess statistically significant violations of invariant item ordering (IIO).

Last, to assess predictive criterion validity, Spearman’s rank correlations between the extracted childhood trauma domains/items and adult health validity measures. To independently assess the individual impact of the extracted items, multivariable logistic and multinomial regression models were conducted including all the trauma domains and further adjusting for baseline age, gender, race/ethnicity, and childhood financial adversity. A complete case analysis was performed. Data cleaning and editing was performed in SAS; scale development and psychometric analyses were performed using RStudio.

## Results

There were 9,340 HRS participants with data sufficient for analysis (79% of LHS respondents; 21% of all HRS participants). Baseline demographic characteristics of the Life History Survey sample are presented by completeness of childhood trauma data in Table 1. Most participants were women (60.7%), White/non-Hispanic (73.3%) and had a high school or general education degree (GED; 54.0%). Compared to those participants with incomplete childhood trauma responses, a greater proportion of our study sample were women (60.7% vs. 58.4%; p-value=0.04), White/non-Hispanic (73.3% vs. 48.4%; p-value<0.01), and more educated (13.0 vs. 22.3% with no degree; p-value<0.01).

**Table 1.** Baseline Demographic Characteristics of the Analytic Sample with Complete versus Incomplete Childhood Trauma Data in the Health and Retirement Study.

|  | Total LHS*<br>Sample<br>(n=11,761) | Complete<br>Trauma Items<br>(n=9,340) | Incomplete<br>Trauma Items<br>(n=2,421) |  |
| --- | --- | --- | --- | --- |
|  | n (%) | n (%) | n (%) | p-value |
| **Gender** |  |  |  | 0.04 |
| Men | 4677 (39.8) | 3669 (39.3) | 1008 (41.6) |  |
| Women | 7084 (60.3) | 5671 (60.7) | 1413 (58.4) |  |
| Missing | 0 | 0 | 0 |  |
| **Race/Ethnicity** |  |  |  | <0.01 |
| White/non-Hispanic | 7983 (68.1) | 6825 (73.3) | 1158 (48.4) |  |
| Black | 2108 (18) | 1430 (15.3) | 678 (28.3) |  |
| Hispanic | 1268 (10.8) | 813 (8.7) | 455 (19.0) |  |
| Other/non-Hispanic | 356 (3) | 255 (2.7) | 101 (4.2) |  |
| Missing | 46 | 17 | 29 |  |
| **Education Level** |  |  |  | <0.01 |
| No Degree | 1750 (14.9) | 1210 (13.0) | 540 (22.3) |  |
| High School/GED | 6175 (52.5) | 5045 (54.0) | 1130 (46.7) |  |
| Some College/Beyond | 3836 (32.6) | 3085 (33.0) | 751 (31.0) |  |
| Missing | 0 | 0 | 0 |  |
| **Age** |  |  |  | 0.10 |
| Mean (SD) | 54.3 (6.7) | 54.2 (6.6) | 54.5 (7.1) |  |
| Median (Range) | 53 (18-87) | 53 (23-86) | 54 (18 - 87) |  |
| Missing | 5 | 0 | 5 |  |

\*LHS=Life History Survey

Self-reported experiences of childhood trauma are reported by race/ethnicity in Table 2. Being separated from a father for ≥6 months (22.8%), death of a parent (21.4%), death of a sibling (18.1%), and problematic substance use by parent(s) (17.5%) were the most reported traumas in the study sample. Less than 2% of the sample reported living in an orphanage, foster family/home or at a boarding school. Compared to White/non-Hispanic adults, a greater proportion of Black and Hispanic/Latino adults reported repeating a year of school (18.7% and 20.4%, respectively vs. 13.1%). Having divorced parents was reported significantly more in Black (22.7%) and Hispanic/Latino (17.7%) than in White adults (12.4%). Nearly a third of Black and Hispanic/Latino participants reported the death of a parent (33.3% and 33.6%, respectively) or of a sibling (29.4% and 32.6%, respectively) before age 18, which was significantly higher than that reported by White adults (17.3% and 12.3%, respectively; p-value<0.01). Over a third (35.9%) of Black adults reported separation from their father and a fifth (21.8%) were separated from their mothers. Whereas nearly half of White adults did not self-report any of the listed ACEs (45.8%), fewer Black (27.5%) and Hispanic/Latino (28.4%) adults reported no ACEs (p-value <0.01).

**Table 2.** Self-reported Childhood Trauma in the Health and Retirement Study by Race/Ethnicity.

|  | White<br>(n=6,825) | Black<br>(n=1,430) | Hispanic<br>(n=813) | Other<br>(n=255) |  |
| --- | --- | --- | --- | --- | --- |
| **Experience** | **n (%)** | **n (%)** | **n (%)** | **n (%)** | **p-value** |
| ***Before the age of 18, did you...*** |  |  |  |  |  |
| Did a year of school over again | 896 (13.1) | 268 (18.7) | 166 (20.4) | 37 (14.5) | <0.01 |
| Got in trouble with the police | 435 (6.4) | 112 (7.8) | 37 (4.6) | 14 (5.5) | 0.02 |
| Parental substance abuse | 1209 (17.7) | 248 (17.3) | 141 (17.3) | 33 (12.9) | 0.27 |
| Physically abused by either parent(s) | 532 (7.8) | 80 (5.6) | 63 (7.8) | 31 (12.2) | <0.01 |
| ***Before the age of 16, did you...*** |  |  |  |  |  |
| Lived in a children's home/orphanage | 99 (1.5) | 19 (1.3) | 13 (1.6) | 6 (2.4) | 0.64 |
| Lived with a foster family/foster home | 106 (1.6) | 31 (2.2) | 10 (1.2) | 2 (0.8) | 0.19 |
| Lived at a boarding school | 116 (1.7) | 24 (1.7) | 13 (1.6) | 6 (2.4) | 0.87 |
| Parents separated/divorce | 843 (12.4) | 324 (22.7) | 144 (17.7) | 34 (13.3) | <0.01 |
| Death of a Parent | 1182 (17.3) | 476 (33.3) | 273 (33.6) | 65 (25.5) | <0.01 |
| Death of a Sibling* | 336 (12.4) | 176 (29.4) | 180 (32.6) | 23 (21.5) | <0.01 |
| Maternal Separation $\geq 6$ months | 634 (9.3) | 311 (21.8) | 191 (23.5) | 52 (20.4) | <0.01 |
| Paternal Separation $\geq 6$ months | 1272 (18.6) | 513 (35.9) | 259 (21.9) | 81 (31.8) | <0.01 |
| Grandparent(s) ever primary caregivers | 322 (4.7) | 217 (15.2) | 107 (13.2) | 26 (10.2) | <0.01 |
| **Total Childhood Trauma Count** |  |  |  |  | <0.01 |
| 0 | 3128 (45.8) | 393 (27.5) | 231 (28.4) | 88 (34.5) |  |
| 1 | 1700 (24.9) | 309 (21.6) | 172 (21.1) | 62 (24.3) |  |
| 2 | 893 (13.1) | 257 (18) | 134 (16.5) | 35 (13.7) |  |
| 3 | 490 (7.2) | 169 (11.8) | 96 (11.8) | 31 (12.2) |  |
| 4 | 300 (4.4) | 145 (10.1) | 86 (10.6) | 20 (7.8) |  |
| $\geq 5$ | 314 (4.6) | 157 (11) | 94 (11.6) | 19 (7.5) | |

Gender differences in self-reported childhood trauma were also noted and presented in Supplemental Table 1. Being separated from a father for ≥6 months (22.8%), death of a parent (21.4%), death of a sibling (18.1%), and problematic substance use by parent(s) (17.5%) were the most reported traumas in the study sample. Less than 2% of the sample reported living in an orphanage, foster family/home or at a boarding school. Compared to men, women were more likely to report parental substance use problems (18.1% vs. 16.4%; p-value=0.03), physical abuse (8.6% vs. 5.9%; p-value<0.01) and parental divorce (15.1% vs. 13.6%; p-value=0.04), and less likely to report repeating a school year (10.8% vs. 20.7%; p-value<0.01), getting in trouble with police (2.1% vs. 13.1%; p-value<0.01), and maternal separation (12.1% vs. 13.7%; p-value=0.03).

Findings from the factor analyses are presented in Supplemental Table 2. With a one factor solution, the domain for family structure disruption emerged, with living in an orphanage, living in a foster home, parental divorce, separation from mother and/or father, and residing with grandparents satisfying the factor loading and communality criterion. A two-factor solution separated out only two items reflecting the death of a parent and/or sibling into a separate domain. With three or more factor solutions, models resulted in factor loadings >1.0 and communalities for items remained ≤ 0.3, indicating the model suggested additional domains but solutions were challenged by poor fit and potentially underrepresented domains. Physical abuse and parental substance use problems did not satisfy the communality criteria for these solutions. Furthermore, the hierarchical omega estimated from evaluating the strength of a potential single common domain underlying the trauma items was *ω*_ℎ_ = 0.32/0.69. These results indicated a lack of support for a single common factor underlying item responses and suggest more than one source of variability in the childhood trauma items. The exploratory factor analysis was inconclusive about the separation of items onto unique factors given the limited number of items available to define potentially unique domains.

Results from the AISP presented two or three hierarchical *Mokken* scales depending on the positive lower bound *c* that was used (*c*=0.4 or *c=*0.3, respectively); item and scale coefficients from the MSA are presented in Table 3. Both solutions clustered (1) living in an orphanage, foster care, parental divorce/separation, separation from mother, separation from father, and living with grandparents as primary caregivers in one domain, and (2) death of a parent and death of a sibling in another domain. Problematic parental substance abuse and physical abuse clustered together in a third domain when a positive lower bound of *c=*0.3 was used. Item scalability coefficients for all the items were positive with *c* ≥ 0.30, and assumptions for monotonicity and non-intersection were met. The first *Mokken* scale, which we termed “*family structure disruption,”* had strong scalability (*H*_*T*_ = 0.55 ≥ 0.5) and the second *Mokken* scale, “*death,”* had moderate scalability (0.4 ≤ *H*_*T*_ = 0.41 < 0.5). The third *Mokken* scale, “*abuse”* had weak scalability (0.3 ≤ *H*_*T*_ = 0.39 < 0.4). Repeating a year of school, getting in trouble with police, and attending boarding school did not meet scalability criteria.

**Table 3.** Item and Scale H Coefficients from *Mokken* Scale Analysis of Childhood Trauma Items in the Health and Retirement Study.

| Domain | Cluster | Item H (SE) | Scale H |
| --- | --- | --- | --- |
| **Disrupted Family Structure Domain** |  |  | 0.55 |
| Lived in a children's home or orphanage | 1 | 0.45 (0.03) |  |
| Lived with a foster family or in a foster home | 1 | 0.52 (0.03) |  |
| Biological/adoptive parents separated/divorce | 1 | 0.46 (0.02) |  |
| Separated from your mother for $\geq 6$ months | 1 | 0.56 (0.01) | |
| Separated from your father for $\geq 6$ months | 1 | 0.72 (0.02) | |
| Grandparent(s) ever your primary caregivers | 1 | 0.47 (0.02) |  |
| **Death Domain** |  |  | 0.41 |
| Death of a Parent | 2 | 0.41 (0.03) |  |
| Death of a Sibling* | 2 | 0.41 (0.03) |  |
| **Abuse Domain** |  |  | 0.39 |
| Physical Abuse | 3 | 0.39 (0.03) |  |
| Parental Substance Abuse | 3 | 0.39 (0.03) |  |

\*Among participants that reported having a sibling

Given the weak scalability of the “abuse” construct, we did not retain parental substance abuse and physical abuse together as a scale; however, given their relevance and mapping onto the existing ACE measures,^1,12^ they were kept as independent items for subsequent predictive inferential analyses. From these findings, external predictive criterion validity analyses were conducted between the identified constructs ([1] family structure disruption, [2] death of parent/sibling, [3] parental substance abuse, and [4] physical abuse) with adult health and socioeconomic indicators. Spearman’s rank coefficients from bivariate associations for each of the four trauma measures were weak overall (Table 4). Spearman’s rank coefficients, *ρ*, between the family structure disruption domain with adult self-rated general health (*ρ* = 0.11), being a current smoker (*ρ* = 0.05), ≥2 drinks per day (*ρ* = 0.05), depression in the last year (*ρ* = 0.07), and BMI category (*ρ* = 0.06) were low. Experiencing the death of a parent and/or sibling in early life had similar correlations, with positive correlations observed with adult self-rated general health (*ρ* = 0.13), being a current smoker (*ρ* = 0.06), ≥2 drinks per day (*ρ* = 0.07), being depressed in the last year (*ρ* = 0.03), and BMI category (*ρ* = 0.06). Spearman rank coefficients for physical abuse and parental substance abuse (evaluated separately) with adult health indicators were comparable to each other and overall weak. Correlations between childhood trauma indicators and childhood financial adversity were also low (i.e., family structure disruption [*ρ* = 0.13], death [*ρ* = 0.13], physical abuse [*ρ* = 0.08], and parental substance abuse [*ρ* = 0.10]). Last, experiences of more childhood trauma were associated with less educational attainment, more so for family structure disruption (*ρ* = −0.08) and parental/sibling death (*ρ* = −0.16).

**Table 4.**
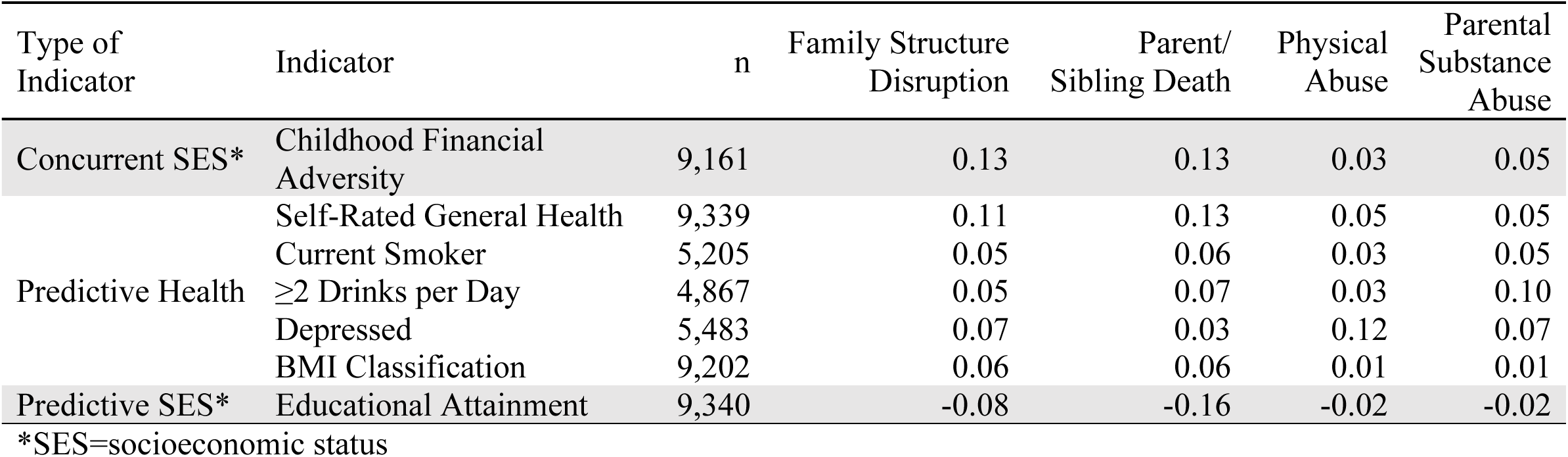
Spearman’s Rank Coefficients from Construct Validity Analyses of Extracted Childhood Trauma Domains in the Health and Retirement Study.

Presented in Table 5 are measures of association from modelling all trauma scales/items together in regressions with the adult health and socioeconomic indicators. Each type of childhood trauma was significantly associated at some level with increased odds of worse (fair or poor versus excellent) self-reported general health. Parental/sibling death was significantly associated with being a current smoker (either parent/sibling vs. neither: OR=1.24, 95% CI: 1.07-1.43; both parent/sibling vs. neither: OR=1.42, 95% CI: 1.06-1.89), while none of the other types of traumas were significantly associated. Parental/sibling death (either parent/sibling vs. neither: OR=1.22, 95% CI: 1.00-1.48; both parent/sibling vs. neither: OR=1.52, 95% CI: 1.04-2.22), and parental drug abuse (yes vs. no: OR= 1.87, 95% CI: 1.54-2.26) were both significantly associated with increased odds of drinking ≥2 alcoholic drinks/day while family structure disruption and physical abuse were not. Increased odds of depression in the past year were observed with family structure disruption (≥3 types vs. 0: OR=1.43, 95% CI:1.14-1.81), physical abuse (yes vs. no: OR= 2.04, 95% CI: 1.64-2.53), and parental substance abuse (yes vs. no: OR= 1.28, 95% CI: 1.08-1.51), but not parental/sibling death. At least one level of family structure disruption, parental/sibling death, and physical abuse were associated with increased odds of being overweight or obese versus normal BMI. Parental/sibling death was associated with significantly lower odds of HS/GED and university versus no degree.

**Table 5.**
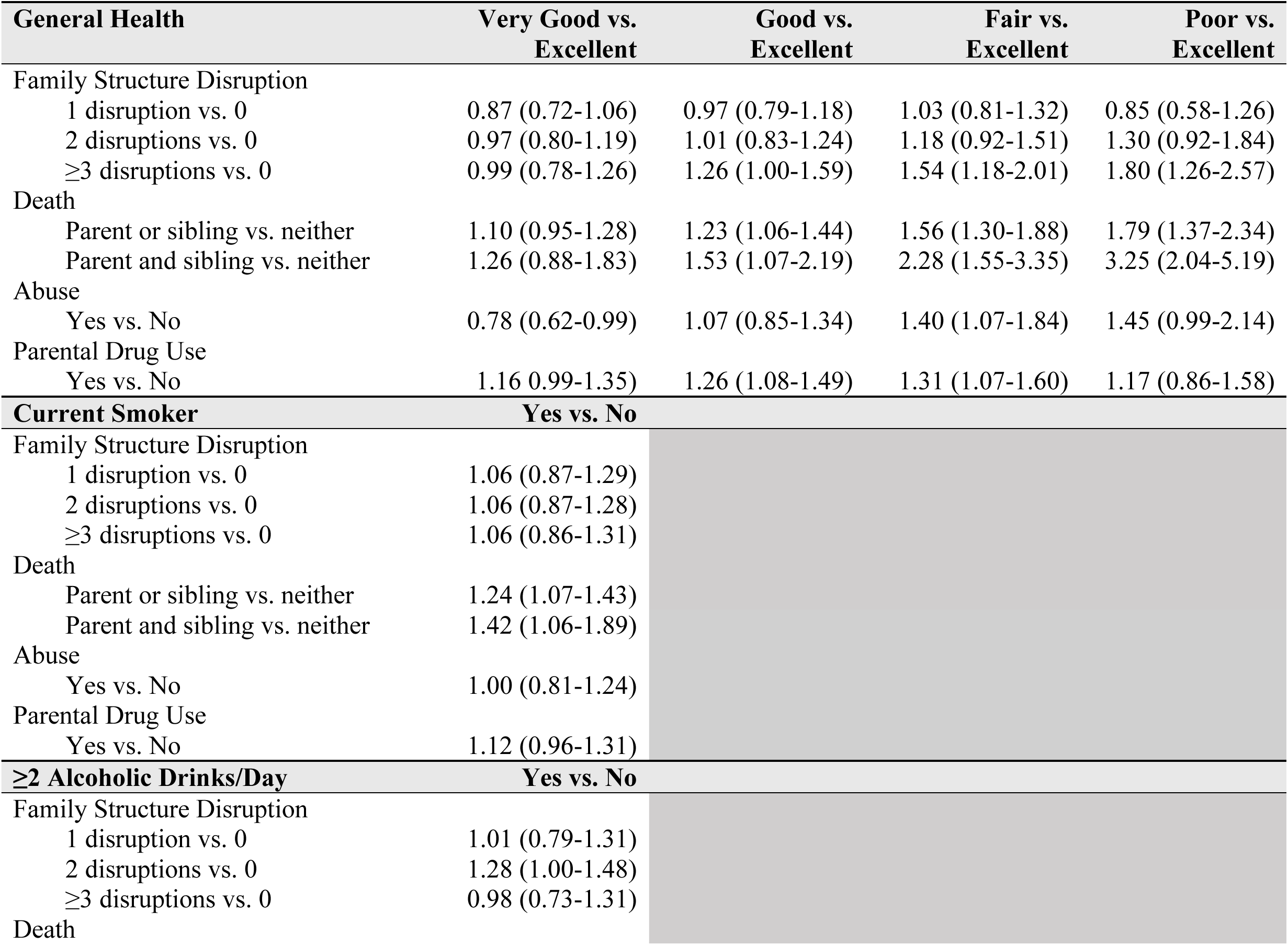
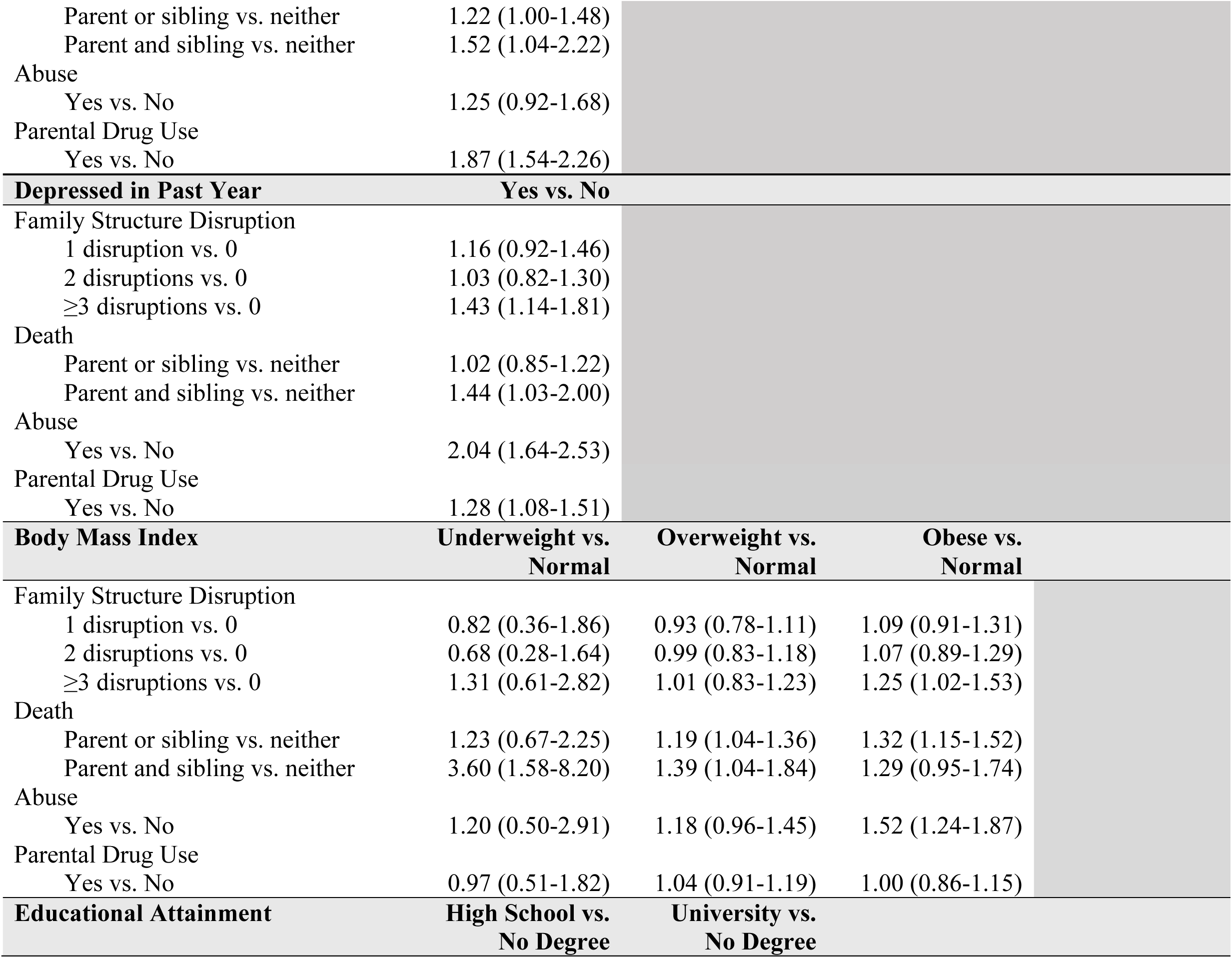
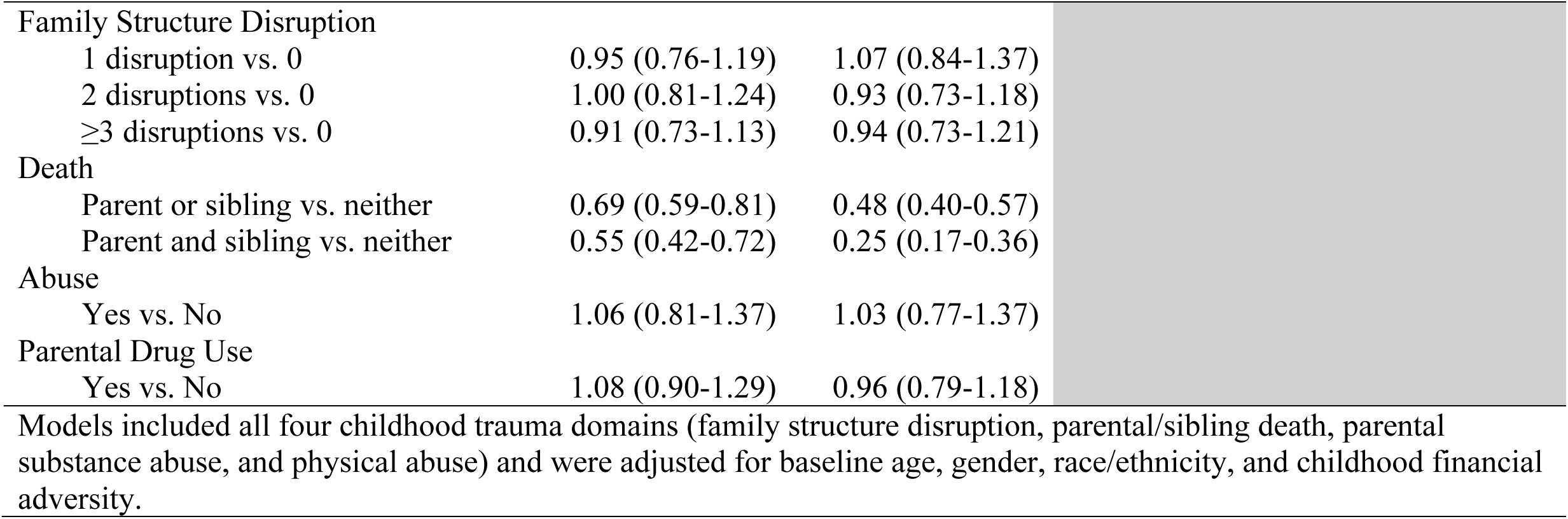
Multivariable Predictive Construct Validity Analyses with Extracted Childhood Trauma Scales and Items.

| **General Health** | **Very Good vs.**<br>**Excellent** | **Good vs.**<br>**Excellent** | **Fair vs.**<br>**Excellent** | **Poor vs.**<br>**Excellent** |
| --- | --- | --- | --- | --- |
| Family Structure Disruption |  |  |  |  |
| 1 disruption vs. 0 | 0.87 (0.72-1.06) | 0.97 (0.79-1.18) | 1.03 (0.81-1.32) | 0.85 (0.58-1.26) |
| 2 disruptions vs. 0 | 0.97 (0.80-1.19) | 1.01 (0.83-1.24) | 1.18 (0.92-1.51) | 1.30 (0.92-1.84) |
| ≥3 disruptions vs. 0 | 0.99 (0.78-1.26) | 1.26 (1.00-1.59) | 1.54 (1.18-2.01) | 1.80 (1.26-2.57) |
| Death |  |  |  |  |
| Parent or sibling vs. neither | 1.10 (0.95-1.28) | 1.23 (1.06-1.44) | 1.56 (1.30-1.88) | 1.79 (1.37-2.34) |
| Parent and sibling vs. neither | 1.26 (0.88-1.83) | 1.53 (1.07-2.19) | 2.28 (1.55-3.35) | 3.25 (2.04-5.19) |
| Abuse |  |  |  |  |
| Yes vs. No | 0.78 (0.62-0.99) | 1.07 (0.85-1.34) | 1.40 (1.07-1.84) | 1.45 (0.99-2.14) |
| Parental Drug Use |  |  |  |  |
| Yes vs. No | 1.16 (0.99-1.35) | 1.26 (1.08-1.49) | 1.31 (1.07-1.60) | 1.17 (0.86-1.58) |
| **Current Smoker** | **Yes vs. No** |  |  |  |
| Family Structure Disruption |  |  |  |  |
| 1 disruption vs. 0 | 1.06 (0.87-1.29) |  |  |  |
| 2 disruptions vs. 0 | 1.06 (0.87-1.28) |  |  |  |
| ≥3 disruptions vs. 0 | 1.06 (0.86-1.31) |  |  |  |
| Death |  |  |  |  |
| Parent or sibling vs. neither | 1.24 (1.07-1.43) |  |  |  |
| Parent and sibling vs. neither | 1.42 (1.06-1.89) |  |  |  |
| Abuse |  |  |  |  |
| Yes vs. No | 1.00 (0.81-1.24) |  |  |  |
| Parental Drug Use |  |  |  |  |
| Yes vs. No | 1.12 (0.96-1.31) |  |  |  |
| **≥2 Alcoholic Drinks/Day** | **Yes vs. No** |  |  |  |
| Family Structure Disruption |  |  |  |  |
| 1 disruption vs. 0 | 1.01 (0.79-1.31) |  |  |  |
| 2 disruptions vs. 0 | 1.28 (1.00-1.48) |  |  |  |
| ≥3 disruptions vs. 0 | 0.98 (0.73-1.31) |  |  |  |
| Death |  |  |  |  |
| Parent or sibling vs. neither | 1.22 (1.00-1.48) |  |  |  |
| Parent and sibling vs. neither | 1.52 (1.04-2.22) |  |  |  |
| Abuse |  |  |  |  |
| Yes vs. No | 1.25 (0.92-1.68) |  |  |  |
| Parental Drug Use |  |  |  |  |
| Yes vs. No | 1.87 (1.54-2.26) |  |  |  |
| **Depressed in Past Year** | **Yes vs. No** |  |  |  |
| Family Structure Disruption |  |  |  |  |
| 1 disruption vs. 0 | 1.16 (0.92-1.46) |  |  |  |
| 2 disruptions vs. 0 | 1.03 (0.82-1.30) |  |  |  |
| ≥3 disruptions vs. 0 | 1.43 (1.14-1.81) |  |  |  |
| Death |  |  |  |  |
| Parent or sibling vs. neither | 1.02 (0.85-1.22) |  |  |  |
| Parent and sibling vs. neither | 1.44 (1.03-2.00) |  |  |  |
| Abuse |  |  |  |  |
| Yes vs. No | 2.04 (1.64-2.53) |  |  |  |
| Parental Drug Use |  |  |  |  |
| Yes vs. No | 1.28 (1.08-1.51) |  |  |  |
| **Body Mass Index** | **Underweight vs. Normal** | **Overweight vs. Normal** | **Obese vs. Normal** |  |
| Family Structure Disruption |  |  |  |  |
| 1 disruption vs. 0 | 0.82 (0.36-1.86) | 0.93 (0.78-1.11) | 1.09 (0.91-1.31) |  |
| 2 disruptions vs. 0 | 0.68 (0.28-1.64) | 0.99 (0.83-1.18) | 1.07 (0.89-1.29) |  |
| ≥3 disruptions vs. 0 | 1.31 (0.61-2.82) | 1.01 (0.83-1.23) | 1.25 (1.02-1.53) |  |
| Death |  |  |  |  |
| Parent or sibling vs. neither | 1.23 (0.67-2.25) | 1.19 (1.04-1.36) | 1.32 (1.15-1.52) |  |
| Parent and sibling vs. neither | 3.60 (1.58-8.20) | 1.39 (1.04-1.84) | 1.29 (0.95-1.74) |  |
| Abuse |  |  |  |  |
| Yes vs. No | 1.20 (0.50-2.91) | 1.18 (0.96-1.45) | 1.52 (1.24-1.87) |  |
| Parental Drug Use |  |  |  |  |
| Yes vs. No | 0.97 (0.51-1.82) | 1.04 (0.91-1.19) | 1.00 (0.86-1.15) |  |
| **Educational Attainment** | **High School vs. No Degree** | **University vs. No Degree** |  |  |

|  |  |  |
| --- | --- | --- |
| Family Structure Disruption |  |  |
| 1 disruption vs. 0 | 0.95 (0.76-1.19) | 1.07 (0.84-1.37) |
| 2 disruptions vs. 0 | 1.00 (0.81-1.24) | 0.93 (0.73-1.18) |
| ≥3 disruptions vs. 0 | 0.91 (0.73-1.13) | 0.94 (0.73-1.21) |
| Death |  |  |
| Parent or sibling vs. neither | 0.69 (0.59-0.81) | 0.48 (0.40-0.57) |
| Parent and sibling vs. neither | 0.55 (0.42-0.72) | 0.25 (0.17-0.36) |
| Abuse |  |  |
| Yes vs. No | 1.06 (0.81-1.37) | 1.03 (0.77-1.37) |
| Parental Drug Use |  |  |
| Yes vs. No | 1.08 (0.90-1.29) | 0.96 (0.79-1.18) |
| Models included all four childhood trauma domains (family structure disruption, parental/sibling death, parental substance abuse, and physical abuse) and were adjusted for baseline age, gender, race/ethnicity, and childhood financial adversity. |  |  |

## Discussion

Results from the current study are informative for future researchers investigating associations between childhood trauma with adverse age-related health outcomes later in the life course. Using available childhood trauma items in the HRS, we identified two moderate-to-strong scalable domains: disruption of family structure and parental/sibling death. Physical abuse and problematic parental substance abuse had weak scalability but are relevant exposures in the childhood adversity literature^1,12^ and should be considered but should be evaluated separately. When the four measures of family structure disruption, death, parental substance abuse, and physical abuse were modeled together, they had significant differential predictive associations with adult health and socioeconomic indicators. These findings underscore the importance of examining distinct domains of childhood trauma instead of a single summed scores to better understand key drivers of adverse health outcomes.

While simple bivariate associations between individual scales/items demonstrated low correlations, the multivariate analyses exploring all the scales/items together and further adjusting for key confounders were notable. Compared to individuals who lost neither, parental/sibling death were associated with worse general health, being a current smoker, and drinking ≥2 drinks/day, and had some of the largest magnitudes of associations with these outcomes. Parental/sibling death may present an early life social determinant of health that begins the accumulation of factors throughout the life course. Our study shows large racial/ethnic differences in the prevalence of parental/sibling death, with nearly a third of Black and Hispanic/Latino adults reporting the death of either or both parent/sibling before age 16 compared to less than one-fifth of White/non-Hispanic adults; this is consistent with racial inequity of parental/sibling death reported in the literature.^13^ Parental/sibling death earlier in the life course presents a risk factor that may result in culminating disadvantage, particularly for Black Americans,^13^ which in combination with weathering from cumulative socioeconomic disadvantage (i.e., the weathering hypothesis)^14–16^ and less access to flexible resources (i.e., the Fundamental Cause Theory)^17^ may precipitate more adverse outcomes in later life. Our analyses lend support to that potential pathway to explain health inequities later in the life course.

After adjustment, both family structure disruption and parental/sibling death were significantly associated with lower educational attainment in our analyses, which has important implications throughout the life course as it pertains to socioeconomic potential, social mobility, and residential neighborhood environments, all of which have an impact on health behaviors and outcomes. This is consistent with other studies that have examined accumulation of adverse childhood experiences with education.^1,4^ The Fundamental Cause Theory posits that access to flexible resources – such as knowledge, power, and wealth – can be used to avoid risk and engage in and with health-promoting environments and resources.^17,18^ As such, differential educational attainment resulting from disruptions in family structure and/or parental/sibling death in early life may lead to a chain of adversity throughout the life course and an accumulation of risk that results in inequities in morbidity and mortality later in life.

In adjusted models, neither physical abuse nor problematic parental substance abuse were significantly associated with being a current smoker or educational attainment; however, reported physical abuse was associated with depression and obesity, while problematic parental substance abuse was associated with ≥2 drinks/day. This underscores the importance of examining distinct types of childhood trauma independently with adverse health outcomes in adulthood to better inform the types of adversity that are driving associations. When childhood trauma measures sum across different types of adversities, important information is lost, and overall associations may be null or largely attenuated by combining the influences of all the adversities in a single metric. Accounting for all the scales/items together in a model while being able to examine and interpret them separately adds to the practicality of using these types of measures to identify/target individuals with tailored prevention and/or risk reductions strategies. This is further demonstrated from the bivariate associations, where we observed low correlations between the extracted childhood trauma indicators with concurrent childhood financial adversity. This is consistent with the concept that exposure to ACEs is orthogonal to childhood socioeconomic status, and that ACEs can occur to any child regardless of their socioeconomic status. It supports the literature that childhood trauma and childhood socioeconomic adversity are distinct exposures that can modify each other and should be examined separately.^5,6^

This study has several strengths. First, these analyses were conducted in a large and diverse U.S.-based aging cohort, and as such, the measure can be used in the context of adverse age-related health outcomes. Additionally, we subjected the items to psychometric and validity analyses; identifying distinct scalable domains of childhood trauma from the existing available HRS items enabled us to understand the specific components that drive associations with adverse outcomes. Our analyses show that associations differ in direction and magnitude depending on the type and level of adversity experienced, which underscores the importance of examining individual trauma constructs/items and not summed scores. Last, we created a measure that was distinct from childhood socioeconomic adversity, which the literature has shown to be a separate type of adversity from the psychosocial trauma captured in ACE measures.^5,6^

Some limitations also warrant discussion. Potentially traumatic items were recalled retrospectively, and in some cases, several decades after the exposure occurred; as such, there may be recall bias especially in older participants. Additionally, the analyses were conducted with the existing set of survey items intended to capture early life adversity in the HRS which differs from the Kaiser Permanente ACE study^1^ and the Center for Disease Control and Prevention (CDC)^12^ACE measures. The HRS did not ask about emotional or sexual abuse, neglect, witnessing domestic violence, or living with a mentally ill parent, among others, but did ask about getting in trouble at school or with police. Thus, we were limited by the number and type of items available to define potentially unique domains, which likely resulted in the inconclusive separation of unique factors. Last, analyses were limited to the Life History Mail Survey subsample who further completed a second questionnaire; about 20% of the subsample was missing items needed for the analyses. The remaining sample was less diverse and more educated than the subsample, and as such, less representative of the U.S. population. Future work should prioritize diverse samples when developing early life trauma/adversity measures.

Despite these limitations, our analyses are valuable in understanding the relationship between different childhood traumas on adverse health and socioeconomic outcomes. We leverage existing data to investigate the impact of a social determinant of health on health-related outcomes and optimize an integral aging cohort, so that we may examine these relationships with later life outcomes. We found that family structure disruption, parental/sibling death, physical abuse, and parental substance abuse are all associated with adverse outcomes and may have implications on adverse age-related outcomes later in the life course.

## Conclusions

Findings from this study help to operationalize early life social determinants of health, such as childhood trauma, in a prominent U.S.-based aging cohort study. The findings facilitate the study of the life course impact of childhood trauma – specifically, family structure disruption, parental/sibling death, physical abuse, and parental substance abuse – on various aging outcomes in later life.

## Data Availability

All data products are publicly available under the aggregated public survey data products for Cross-Wave Childhood Health And Family Aggregated Data. https://hrsdata.isr.umich.edu/data-products/public-survey-data

https://hrsdata.isr.umich.edu/data-products/cross-wave-lifecourse-experiences-adversity-and-trauma-aggregated-data

## Notes

### Competing Interest Statement

The authors have declared no competing interest.

### Funding Statement

This work was supported by the National Institute of Health (grant numbers: T32AG058529, F99AG083275, K00AG083275).

### Author Declarations

The study used only publicly available survey data originally located at: https://hrsdata.isr.umich.edu/data-products/cross-wave-lifecourse-experiences-adversity-and-trauma-aggregated-data.

