## Supplemental Table 1, Supplemental Table 2 for "Measuring Childhood Trauma among Adults in the Health and Retirement Study"

Supplemental Table 1. Self-reported Childhood Trauma in the Health and Retirement Study by Gender

| | Total (n=9,340) | Men (n=3,669) | Women (n=5,671) | |
|---|---|---|---|---|
| **Experience** | n (%) | n (%) | n (%) | p-value |
| ***Before the age of 18, did you…*** | | | | |
| Did a year of school over again | 1370 (14.7) | 758 (20.7) | 612 (10.8) | <0.01 |
| Got in trouble with the police | 589 (6.4) | 480 (13.1) | 118 (2.1) | <0.01 |
| Parental substance abuse | 1631 (17.5) | 602 (16.4) | 1029 (18.1) | 0.03 |
| Physically abused by either parent(s) | 707 (7.6) | 218 (5.9) | 489 (8.6) | <0.01 |
| ***Before the age of 16, did you…*** | | | | |
| Lived in a children's home/orphanage | 137 (1.5) | 63 (1.7) | 74 (1.3) | 0.11 |
| Lived with a foster family/foster home | 149 (1.6) | 58 (1.6) | 91 (1.6) | 0.93 |
| Lived at a boarding school | 159 (1.7) | 73 (2) | 86 (1.5) | 0.08 |
| Parents separated/divorce | 1351 (14.5) | 497 (13.6) | 854 (15.1) | 0.04 |
| Death of a Parent | 2000 (21.4) | 804 (21.9) | 1196 (21.1) | 0.34 |
| Death of a Sibling* | 719 (18.1) | 288 (18.7) | 431 (17.7) | 0.40 |
| Maternal Separation ≥6 months | 1191 (12.8) | 503 (13.7) | 688 (12.1) | 0.03 |
| Paternal Separation ≥6 months | 2131 (22.8) | 832 (22.7) | 1299 (22.9) | 0.8 |
| Grandparent(s) ever primary caregivers | 673 (7.1) | 267 (7.3) | 406 (7.2) | 0.83 |
| Total Childhood Trauma Count | | | | <0.01 |
| 0 | 3847 (41.2) | 1380 (37.6) | 2467 (43.5) | |
| 1 | 2245 (24) | 923 (25.2) | 1322 (23.3) | |
| 2 | 1323 (14.2) | 565 (14.5) | 758 (13.4) | |
| 3 | 787 (8.4) | 306 (8.3) | 481 (8.5) | |
| 4 | 552 (5.9) | 215 (5.9) | 337 (5.9) | |
| ≥5 | 586 (6.3) | 280 (7.6) | 306 (5.4) | |

Supplemental Table 2. Results from Childhood Trauma Item Factor Analysis in the Health and Retirement Study - Loading Factors and Communalities from One-, Two-, and Three-Factor Solutions

| | One Factor | | Two Factor | | | Three Factor | | | |
|---|---|---|---|---|---|---|---|---|---|
| | FA1 | $h^2$ | FA1 | FA2 | $h^2$ | FA1 | FA2 | FA3 | $h^2$ |
| Parental Substance Abuse | 0.31 | 0.10 | **0.46** | -0.19 | 0.17 | 0.09 | 0.04 | **0.48** | 0.25 |
| Ever physically abused by either of your parents | 0.39 | 0.15 | **0.55** | -0.2 | 0.25 | 0.06 | 0.15 | **0.58** | **0.38** |
| Ever lived in a children's home or orphanage | **0.69** | **0.47** | **0.64** | 0.11 | **0.47** | -0.29 | **1.11** | 0.26 | **1.01** |
| Ever lived with a foster family or in a foster home | **0.74** | **0.54** | **0.66** | 0.15 | **0.54** | 0.03 | **0.71** | 0.23 | **0.60** |
| Ever lived at a boarding school | **0.43** | 0.19 | 0.33 | 0.17 | 0.18 | -0.01 | **0.51** | -0.06 | 0.26 |
| Biological/adoptive parents were separated/divorce | **0.64** | **0.41** | **0.73** | -0.08 | 0.49 | **0.74** | -0.14 | **0.43** | **0.64** |
| Death of a Parent | **0.46** | 0.21 | -0.04 | **0.83** | **0.66** | 0.36 | 0.32 | -0.29 | **0.44** |
| Death of a Sibling | 0.31 | 0.09 | -0.12 | **0.68** | **0.41** | 0.35 | 0.1 | -0.35 | 0.29 |
| Separated from your mother for ≥6 months | **0.96** | **0.93** | **0.73** | 0.38 | **0.92** | **0.55** | **0.51** | 0.1 | **0.94** |
| Separated from your father for ≥6 months | **0.91** | **0.82** | **0.81** | 0.19 | **0.82** | **0.81** | 0.13 | 0.27 | **0.91** |
| Grandparent(s) ever your primary caregivers | **0.57** | **0.33** | **0.49** | 0.15 | **0.33** | **0.87** | -0.19 | 0.06 | **0.58** |
